# COVID19 vaccine type and humoral immune response in patients receiving dialysis

**DOI:** 10.1101/2021.08.02.21261516

**Authors:** Pablo Garcia, Shuchi Anand, Jialin Han, Maria Montez-Rath, Sumi Sun, Tiffany Shang, Julie Parsonnet, Glenn M Chertow, Brigitte Schiller, Graham Abra

**Affiliations:** Department of Medicine (Nephrology), Stanford University; Satellite Healthcare; Departments of Medicine (Infectious Diseases and Geographic Medicine), and Epidemiology and Population Health, Stanford University; Departments of Medicine (Nephrology), and Epidemiology and Population Health, Stanford University

**Keywords:** dialysis, covid-19, vaccine, SARS-CoV-2, ESRD

## Abstract

**Background:** Patients on dialysis vaccinated with the attenuated adenovirus SARS-CoV-2 vaccine might mount an impaired response to vaccination.

**Methods:** We evaluated the humoral vaccination response among 2,099 fully vaccinated patients receiving dialysis. We used commercially available assays (Siemens) to assess prevalence of no response or diminished response to COVID-19 vaccination by vaccine type. We defined “no seroconversion” as lack of change from negative to positive in total RBD Ig antibody, no detectable response on semiquantitative RBD IgG antibody (index value <1) as “no RBD IgG response”, and a semiquantitative RBD IgG index value <10 as “diminished RBD IgG response”

**Results:** Of the 2,099 fully vaccinated patients on dialysis, the proportion receiving the mRNA1273, BNT162b2, and Ad26.COV2.S were 62% (n=1316), 20% (n=416) and 18% (n=367), respectively. A third (33.3%) of patients receiving the attenuated adenovirus Ad26.COV2.S vaccine failed to seroconvert and an additional 36% had no detectable or diminished IgG response even 28-60 days post vaccination.

**Conclusion:** One in three fully vaccinated patients receiving dialysis had evidence of an impaired immune response to the attenuated adenovirus Ad26.COV2.S vaccine.

---

Patients receiving dialysis may have a less robust antibody response to COVID-19 vaccination, yet have a 10-15 fold higher risk for COVID-19-associated mortality than the general population^1, 2^. We previously raised concern about diminished vaccine responses to the attenuated adenovirus SARS-CoV-2 vaccine compared to m-RNA vaccines among patients on dialysis, although the number of patients receiving the attenuated adenovirus vaccine was small^2^. Here we report qualitative and semi-quantitative receptor-binding domain (RBD) antibody responses by vaccine type and dialysis modality in 2,099 patients receiving dialysis.

In partnership with a non-profit dialysis provider that serves patients undergoing dialysis in four states (California, Texas, Tennessee, and New Jersey), we evaluated the humoral vaccination response among patients receiving dialysis. From 6,022 patients receiving dialysis, we identified 2,099 patients who had been fully vaccinated and whom we had tested for SARS-CoV-2 antibody before and at least 14 days after vaccination. The dialysis provider made no recommendations regarding type of vaccine, and administration was based on supply availability. More patients residing in the South were offered the attenuated adenovirus vaccine due to timing (i.e., facilities in the West and East received vaccines at earlier time points, when attenuated adenovirus vaccines were not yet available).

We tested antibody response using the Siemens’ total receptor-binding domain (RBD) Ig assay, which measures IgG and IgM antibodies^3^. Among those with a total RBD response, we quantified their antibody response using one of two semiquantitative Siemens RBD IgG assays^4, 5^. We defined “no seroconversion” as lack of change from negative to positive in total RBD Ig antibody, no detectable response on semiquantitative RBD IgG antibody (index value <1) as “no RBD IgG response”, and a semiquantitative RBD IgG index value <10 as “diminished RBD IgG response” (Supplementary Methods delineates rationale for index value cut points).

Of the 2,099 fully vaccinated patients on dialysis, the proportion receiving the mRNA1273 (Moderna mRNA), BNT162b2 (Pfizer mRNA), and Ad26.COV2.S (Jonhson and Johnson, attenuated adenovirus) were 62% (n=1316), 20% (n=416) and 18% (n=367), respectively (**Supplement Table 1**). Patients vaccinated with the Ad26.COV2.S vaccine were younger, more likely to be non-Hispanic Black, to have prior evidence of SARS-CoV-2 infection, and to reside in the South. Seroconversion to Ad26.COV2.S vaccine occurred later than to the mRNA vaccines (**Table 1**). Whether assessed during the day 14-28 window post vaccination or in the day 28-60 window post vaccination, however, patients receiving the Ad26.COV2.S vaccine had higher likelhood of no seroconversion and no detectable or diminished IgG response compared with those receiving the mRNA vaccines (**Table 1**). Patients vaccinated with BNT162b2 had higher prevalence of no detectable or diminished IgG response, compared with patients vaccinated with mRNA1273.

**Table 1.** Prevalence of absent or diminished response among fully vaccinated individuals by vaccine type, dialysis modality and overall, between 14 and 28 days and between 28 and 60 days after completion of vaccine*

|  | Fully Vaccinated Cohort<br>Days 14 and 28<br>N=1583 |  | Fully Vaccinated Cohort<br>Days 28 and 60<br>N=1468 |  | *Data are percentage (95% CI) obtained at least 14-28 days, or >28-60 days after two doses of either mRNA1273 or BNT162b2 vaccines and a single dose of Ad26.COV |
| --- | --- | --- | --- | --- | --- |
|  | No seroconversion on total RBD Ig | No detectable or diminished response on RBD IgG ** | No seroconversion on total RBD Ig | No detectable or diminished response on RBD IgG ‡ |  |
| **Vaccine type** |  |  |  |  |  |
| mRNA1273 | 2.6 (1.7, 3.7) | 5.2 (4.0, 6.8) | 2.1 (1.3, 3.3) | 9.4 (7.5, 11.6) |  |
| BNT162b2 | 4.3 (2.5, 7.4) | 11.4 (8.1, 15.7) | 4.0 (2.4, 6.6) | 22.7 (18.6, 27.5) |  |
| Ad26.COV2.S | 58.4 (52.6, 64.0) | 16.8 (12.9, 21.6) | 33.3 (27.3, 40.0) | 36.0 (29.2, 43.5) |  |
| **Dialysis modality** |  |  |  |  |  |
| Home | 13.7 (9.5, 19.3) | 8.4 (5.2, 13.3) | 9.0 (5.4, 14.6) | 14.7 (10.0, 21.2) |  |
| In-center | 12.8 (11.1, 14.7) | 8.5 (7.1, 10.1) | 6.8 (5.5, 8.3) | 16.5 (14.5, 18.7) |  |
| **Vaccine type by dialysis modality** |  |  |  |  |  |
| mRNA1273 |  |  |  |  |  |
| Home | 4.3 (1.8, 10.0) | 1.7 (0.4, 6.7) | 2 (0.5, 7.8) | 7.1 (3.4, 14.2) |  |
| In-center | 2.3 (1.5, 3.5) | 5.7 (4.3, 7.5) | 2.1 (1.3, 3.4) | 9.7 (7.7, 12.1) |  |
| BNT162b2 |  |  |  |  |  |
| Home | 2.6 (0.4, 16.1) | 12.8 (5.4, 27.3) | 5.4 (1.4, 19.2) | 18.9 (9.3, 34.7) |  |
| In-center | 4.6 (2.6, 8.1) | 11.1 (7.7, 15.8) | 3.8 (2.2, 6.5) | 23.2 (18.8, 28.3) |  |
| Ad26.COV2.S |  |  |  |  |  |
| Home | 55.6 (39.3, 70.7) | 25 (13.5, 41.5) | 47.6 (27.8, 68.2) | 42.9 (24.0, 64.0) |  |
| In-center | 58.8 (52.6, 64.7) | 15.6 (11.6, 20.6) | 31.7 (25.5, 38.7) | 35.1 (27.9, 43.0) |  |
| **Overall** | **13.0 (11.4, 14.7)** | **8.5 (7.2, 10.0)** | **7.0 (5.8, 8.4)** | **16.3 (14.4, 18.4)** |  |

2.S vaccine. Diminished classification refers to semiquantitative IgG index value < 10.
\*\*Total N = 1532 due to 51 missing semiquantitative IgG result
‡Total N = 1326 due to 142 missing semiquantitative IgG result

Patients receiving home dialysis with either peritoneal dialysis or home hemodialysis had similar prevalence of no seroconversion and no detectable or diminished IgG response, compared with patients receiving in-center dialysis (**Table 1**). There was no difference in response rates by vaccine type among the home versus in-center populations.

In this large cohort of patients receiving dialysis, prevalence of no response or diminished response to COVID-19 vaccination varied by vaccine type, with a third of patients receiving the attenuated adenovirus Ad26.COV2.S vaccine failing to seroconvert and an additional third having no detectable or diminished IgG response even 28-60 days post vaccination. Although patients receiving home dialysis represent a cohort with better health and functional status, we observed no difference in response rates by dialysis modality, with similar immunogenicity of the three vaccines in circulation in the US among patients on in-center versus home dialysis.

Limitations of this study include the concentration of patients in four states and non-random allocation of vaccine type, although the cohort receiving the attenuated adenovirus vaccine were younger and more likely to have had prior SARS-CoV-2 infection, both factors that likely could have improved response. Although a detectable serum antibody response is often equated with immunity, this relationship is not absolute. We were unable to test for cellular immunity or the presence of memory B cells.

Follow-up of vaccinated patients with end-stage kidney disease for clinical COVID-19 will provide a better estimate of vaccine efficacy by vaccine type. Higher rates of spike protein seroconversion after a third dose of mRNA vaccine were recently reported among solid-transplant organ recipients^6^, and in preliminary data among patients on dialysis^7^. Whether additional vaccination doses or change in vaccine type are necessary for a subset of patients receiving dialysis who have failed to seroconvert or have demonstrated weak immune response in the early period post vaccination requires further investigation.

## Data Availability

Requests for data will be reviewed by the authors, and data made available on a case-by-case basis.

