## Supplement Table 1 for "COVID19 vaccine type and humoral immune response in patients receiving dialysis"

**Table of Contents**

**Supplemental Methods** **p2**

**Supplemental Table 1** Characteristics of patients on dialysis by vaccine type **p3**

### Supplemental Methods

#### Assay Characteristics

The Siemens total RBD Ig assay measures IgG and IgM antibodies. This assay is reported by the manufacture to have 100% sensitivity and 99.8% specificity for tests performed ≥14 days after a positive reverse transcriptase polymerase chain reaction test[1]; it has been validated independently with similar performance characteristics[2, 3]. We used one of two Siemens RBD IgG assays, which are semiquantitative two-step sandwich indirect chemiluminescent assay with a manufacturer reported 95.6% (95% CI: 92.2-97.8%) sensitivity and 99.9% (95% CI 99.6-99.9%) specificity for tests performed ≥21 days post positive reverse transcriptase polymerase chain reaction test. An index value ≥1.0 is considered reactive. The two assays are compatible using a conversion factor, but one has a larger maximal index value range (>44 versus > 150).

We classified responses as absent total RBD Ig antibody, absent semiquantitative IgG antibody (index value <1), diminished antibody (semiquantitative IgG index value <10), or medium to high antibody (≥10)[4]. We chose index value ≥10 as a cut-point based on a Siemens study (n=74) evaluating correlation with plaque reduction neutralization test which reported index values ≥10 had a positive predictive value of 100% for plaque reduction neutralization test50 >1:80.

**Supplemental Table 1** Characteristics of patients on dialysis by vaccine type

| Patient characteristics | Overall N=2099 | mRNA1273 N=1316 | BNT162b2 N=416 | Ad26.COV2.S N=367 |
|---|---|---|---|---|
| **Age** | | | | |
| 18-44 | 180 (8) | 87 (7) | 36 (9) | 57 (16) |
| 45-64 | 685 (33) | 365 (28) | 106 (25) | 214 (58) |
| 65-79 | 885 (42) | 606 (46) | 195 (47) | 84 (23) |
| ≥80 | 349 (17) | 258 (20) | 79 (19) | 12 (3) |
| **Gender** | | | | |
| M | 1277 (61) | 792 (60) | 264 (63) | 221 (60) |
| F | 822 (39) | 524 (40) | 152 (37) | 146 (40) |
| **Race and Ethnicity** | | | | |
| Hispanic | 721 (34) | 406 (31) | 165 (40) | 150 (41) |
| Non-Hispanic white | 522 (25) | 316 (24) | 109 (26) | 97 (26) |
| Non-Hispanic Black | 209 (10) | 81 (6) | 28 (7) | 100 (27) |
| Asian | 312 (15) | 263 (20) | 42 (10) | 7 (2) |
| Other | 61 (3) | 49 (4) | 9 (2) | 3 (1) |
| Missing | 274 (13) | 201 (15) | 63 (15) | 10 (3) |
| **Geographic region** | | | | |
| South | 464 (22) | 73 (6) | 38 (9) | 353 (96) |
| West | 1635 (78) | 1243 (94) | 378 (91) | 14 (4) |
| **Dialysis Modality** | | | | |
| Hemodialysis | 1176 (84) | 1111 (85) | 354 (85) | 311 (85) |
| CCPD | 266 (13) | 176 (13) | 49 (12) | 41 (11) |
| CAPD | 25 (1) | 18 (1) | 2 (1) | 5 (1) |
| Home Hemodialysis | 32 (2) | 11 (1) | 11 (2) | 10 (3) |
| **ESKD Vintage (years)** | | | | |
| < 2 | 799 (38) | 506 (38) | 173 (42) | 120 (33) |
| 2 – 5 | 726 (35) | 455 (35) | 143 (34) | 128 (35) |
| ≥ 5 | 574 (27) | 355 (27) | 100 (24) | 119 (32) |
| **Diabetes** | | | | |
| Yes | 1350 (64) | 848 (64) | 258 (62) | 244 (66) |
| No | 749 (36) | 464 (36) | 158 (38) | 123 (34) |
| **RBD antibody status prior to vaccine^** | | | | |
| Seropositive | 696 (33) | 361 (27) | 141 (34) | 194 (53) |
| Seronegative | 1403 (67) | 955 (73) | 275 (66) | 173 (47) |

Abbreviations: CCPD, continuous cycler-assisted peritoneal dialysis; CAPD, continuous automated peritoneal dialysis; ESKD, end-stage kidney disease; RBD, receptor-binding domain;
^seropositive prior to vaccination indicates likely SARS-CoV-2 infection prior to vaccination
